# Local-Global breakdown of Cortical Similarity Networks in Anorexia Nervosa

**DOI:** 10.1101/2025.10.29.25339093

**Authors:** Massimiliano Facca, Valentina Meregalli, Sofia Gentili, Alessandra Bertoldo, Renzo Manara, Angela Favaro, Enrico Collantoni

**Author notes:** **Corresponding author:** Enrico Collantoni –. Address: Department of Neuroscience, University of Padua, Via Giustiniani, 2 - 35128 Padova.

## Abstract

Anorexia nervosa (AN) profoundly alters brain structure, yet the large-scale organization of the cortex under severe nutritional stress remains poorly understood. In this study, we map cortical architecture in 50 patients with AN and 40 healthy controls using Morphometric INverse Divergence (MIND), a multivariate framework integrating multiple morphometric features into cortical similarity networks. Patients showed a marked global reduction in morphometric similarity that scaled with nutritional status, with lower body-mass index associated with weaker global and network-level connectivity. Disruptions selectively involved transmodal systems including limbic, default mode, frontoparietal and dorsal attention, whereas primary sensorimotor and visual cortices were relatively preserved. Region-level effects followed the topology of the structural connectome, and network-based statistics revealed weakened long-range coupling between attentional and limbic–prefrontal circuits. Spatial correspondence with cortical gradients of serotonergic, dopaminergic, μ-opioid and CB1 receptor densities suggests that malnutrition compromises cortical integrity along neurochemically defined axes of vulnerability.

## Introduction

Anorexia nervosa (AN) is a severe psychiatric disorder marked by persistent energy restriction, intense fear of weight gain, and disturbance in body image, with wide cognitive–affective and medical sequelae (DSM-5; Zipfel et al. 2015). Onset concentrates in adolescence, a period of intense cortical remodeling and high metabolic demand, when nutritional deprivation can critically constrain neurodevelopment (Fuhrmann et al. 2015). This timing places AN at the interface of maturational vulnerability and neuroprogression, suggesting that brain alterations may reflect both early developmental constraints and later disorder related processes (Schmidt 2025).

Large-scale neuroimaging studies have demonstrated widespread cortical gray matter reductions in acute AN, among the most extensive reported in psychiatry. These abnormalities, particularly in cortical thickness, tend to normalize with partial weight recovery, implicating nutritional state while not excluding longer-lived alterations (Walton et al. 2022). Beyond thickness, longitudinal and cross-sectional studies indicate abnormalities in other cortical measures such as gyrification, sulcal morphology, and surface area, with partial normalization after refeeding and age-dependent constraints on recovery (Collantoni et al. 2021 Kaufmann et al. 2020). The close temporal association between weight restoration and rapid cortical re-thickening underscores the critical role of nutritional state and metabolic stressors in shaping cortical architecture. Yet, the heterogeneity observed across distinct morphometric indices suggests that these changes arise from multiple, partially dissociable biological processes rather than a single reversible effect of malnutrition (King et al. 2018). While acute undernutrition likely drives much of the cortical alteration observed in AN, pre-existing neurobiological susceptibilities possibly related to maturational timing, metabolic efficiency, or neurotransmitter regulation, may influence both the emergence and persistence of these alterations. Such mechanisms could contribute to treatment resistance and to the long-term course of the disorder, linking transient metabolic effects with enduring aspects of disease vulnerability (Collantoni et al. 2019; Ehrlich et al. 2025). From a systems perspective, the human cortex is organized into a hierarchical network of regions that differ in metabolic cost and connectivity demand. High-degree hubs, especially those integrating transmodal networks, are metabolically expensive and selectively vulnerable across neurological and psychiatric disorders (Bullmore & Sporns 2009). Embedding disease-related maps in normative connectomes consistently reveals hub susceptibility, driven by high activity, wiring cost, and molecular burden (Crossley et al. 2014). Thus, interpreting AN-related cortical changes as network-constrained deviations rather than isolated regional effects may better capture their underlying biology. This perspective aligns with etiological evidence indicating that AN arises from an interplay between metabolic and psychiatric risk factors. Genome-wide association studies have revealed a metabo-psychiatric genetic architecture, linking loci involved in energy homeostasis and behavioral regulation, suggesting that metabolically demanding hub networks may represent preferential sites of cortical vulnerability (Watson et al. 2019). Complementary receptor-imaging and pharmacological findings indicate involvement of serotonergic, dopaminergic, opioid, and endocannabinoid systems as molecular substrates of vulnerability in AN (Bailer et al. 2010; Miranda-Olivos et al. 2023; Pak et al. 2025). Moreover, elevated neurofilament light levels during acute illness indicate active neuroaxonal stress, consistent with a metabolically sensitive and connectome-dependent pathophysiology (Hellerhoff et al. 2023).

Despite this convergent evidence, most structural MRI studies of AN have relied on single-feature analyses (e.g., thickness, gyrification, or surface area), which fragment the signal across modalities, obscure shared variance, and constrain biological interpretation. Here we adopt Morphometric INverse Divergence (MIND), a multivariate framework that models each cortical region by its vertex-wise distribution of multiple features and quantifies interregional similarity from the divergence between these distributions. In benchmark datasets, MIND shows a closer correspondence to cytoarchitectonics, tract-tracing connectivity, and cortical gene co-expression, thereby supporting multiscale inference from standard structural MRI (Sebenius et al., 2023). Applied to AN, this approach could offer a connectome-grounded lens for situating case–control differences within cortical network architecture and for examining their alignment with underlying molecular organization.

In this study, we applied the MIND framework to structural MRI data from individuals with AN and healthy controls. We hypothesized that cortical morphometric similarity would be reduced in AN, reflecting (i) the severity of malnutrition, (ii) the preferential involvement of metabolically demanding hub systems, and (iii) spatial correspondence between regional MIND alterations and cortical gradients of serotonergic, dopaminergic, opioid, and endocannabinoid receptor or transporter densities - with a priori expectation of stronger correspondence for appetite- and reward-related systems, which are central to AN pathophysiology.

## Results

This study included a cohort of 90 participants (N = 50 with AN and N = 40 healthy controls). Patients with AN had a mean age of 20.4 years (SD=5.68) and a mean BMI of 15.69 (SD=0.59). Healthy controls had a mean age of 25.3 years (SD=3.17) and a mean BMI of 20.23 (SD=1.82). For each individual, a structural similarity matrix was derived in two steps. First, cortical surface reconstruction was performed using FreeSurfer’s *recon-all* pipeline (Fischl 2012). Second, we applied the Morphometric INverse Divergence (MIND) approach to compute a matrix capturing pairwise similarity between regions of interest (ROIs) based on FreeSurfer-derived morphometric features (see Methods for details). We employed the Schaefer 100 regions 7 Networks atlas (Schaefer et al. 2018), resulting in a 100 × 100 symmetric MIND matrix per subject (Figure 1a,b). Age was included as a covariate in all analyses to account for the slight age difference between groups, together with Total Intracranial Volume (TIV) to control for differences in brain size.

**Figure 1.**
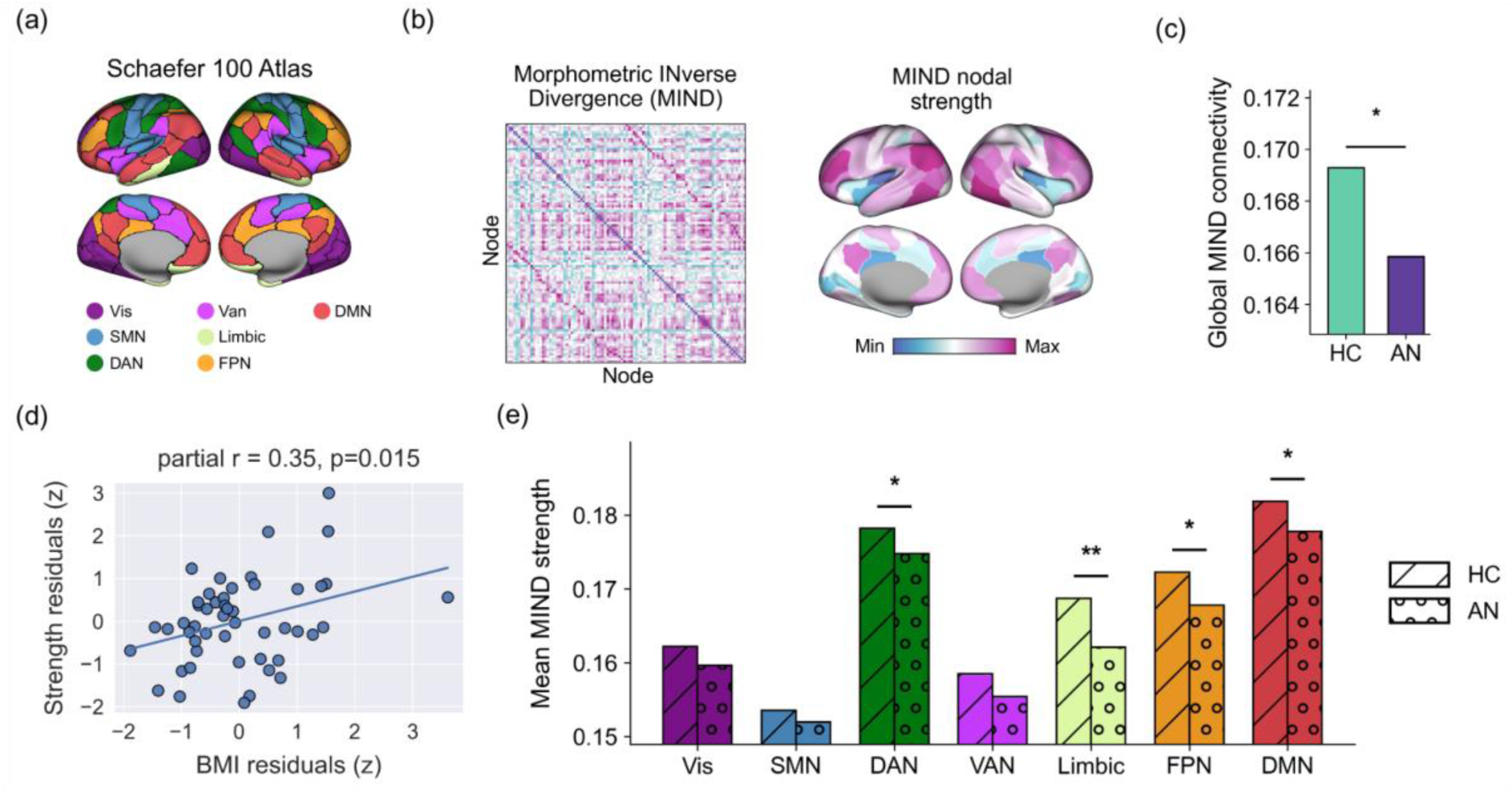
Overview of the analysis and global and network specific alterations of MIND connectivity in AN. (a) All analyses were performed using the Schaefer 100 regions 7 networks atlas (Schaefer et al. 2018). (b) For each subject, a structural covariance matrix was generated using the Morphometric INverse Divergence (MIND) approach and the chosen parcellation. The resulting symmetric matrix can be examined at global, network, and region-specific levels. The right panel shows regional connectivity strength in the healthy control group. (c) Global connectivity differences between patients with anorexia nervosa (AN) and healthy controls (HC). Patients with AN exhibited reduced global structural covariance compared with HC. (d) Partial correlation between body-mass index (BMI) and global MIND strength in the AN group. Both global strength and BMI were residualized for age and total intracranial volume. (e) Network-specific MIND strength differences between AN and HC. * indicates statistical significance (*p < 0.05, **p < 0.01; FDR-corrected).

### 1.1 Global loss of connectivity strength in AN

We first compared global MIND connectivity between groups. Global connectivity strength was defined as the mean of all edges in each subject’s MIND network. After adjusting for age and TIV, individuals with AN showed reduced global connectivity strength compared with healthy controls (*t* = 2.10; one-sided *p* = 0.019, Figure 1c). Within the AN group, global connectivity strength was positively associated with body mass index (BMI) after controlling for age and TIV (*t* = 2.47; *p* = 0.017; partial *r* = 0.35), indicating that lower BMI was linked to reduced global MIND connectivity (Figure 1d). No significant relationship between BMI and global strength was observed in healthy controls (*t* = −1.28; *p* = 0.21; partial *r* = −0.21).

### 1.2 Network-specific connectivity alterations in AN

We next examined connectivity strength across canonical Resting-State Networks (RSNs, Yeo 2011) using the same GLM framework. After adjusting for age and TIV, individuals with AN showed weaker structural covariance than healthy controls in several higher-order networks. Significant differences (FDR-corrected *p* < 0.05) were observed in the limbic (*t* = 3.15; *p* = 0.008), FPN (*t* = 2.70; *p* = 0.015), DMN (*t* = 2.37; *p* = 0.024) and DAN (*t* = 2.15; *p* = 0.03) networks (Figure 1e). In contrast, the salience/ventral attention network showed only a trend towards significance (*t* = 1.77; FDR-corrected *p* = 0.06), and no significant differences were detected in the SMN or Vis networks (all FDR-corrected *p* > 0.20). These findings indicate that connectivity loss was mainly restricted to transmodal systems, reflecting a selective disruption of higher-order networks in AN. Exploratory analyses revealed positive partial correlations between BMI and mean network strength (controlling for age and TIV) in several RSNs, with the largest effects in FPN (partial *r* = 0.36; *p* = 0.012), DMN (partial *r* = 0.32; *p* = 0.026) and DAN (partial *r* = 0.31; *p* = 0.033) networks. Although none of these associations survived FDR correction (all *p* > 0.05), the pattern was consistent with preferential involvement of transmodal systems.

### 1.3 Region-specific connectivity strength alterations in AN

We next investigated region-wise differences in MIND connectivity between AN and healthy controls using the same GLM framework. Before multiple-comparison correction, reductions of node strength in the AN group were widespread across the cortex. Ten regions survived FDR correction (all *p* < 0.05; FDR-corrected), predominantly located in transmodal systems (Figure 2). This pattern mirrored the global and network-level findings, indicating that the loss of MIND connectivity in patients with AN is particularly pronounced in DMN and limbic areas (See Supplementary Table T1 for the full list of significant regions). We then examined region-wise associations between BMI and node strength within the AN group, controlling for age and TIV. After FDR correction, significant positive correlations emerged in regions belonging to the VAN, Vis and DMN networks, with partial correlations ranging from *r* = 0.43 to 0.52 (all *p* < 0.05; FDR-corrected, Supplementary Table T2 for the full list of significant regions). These results indicate that lower BMI, indicative of greater illness severity, was associated with stronger reductions of MIND connectivity at the regional level (Figure 3).

**Figure 2.**
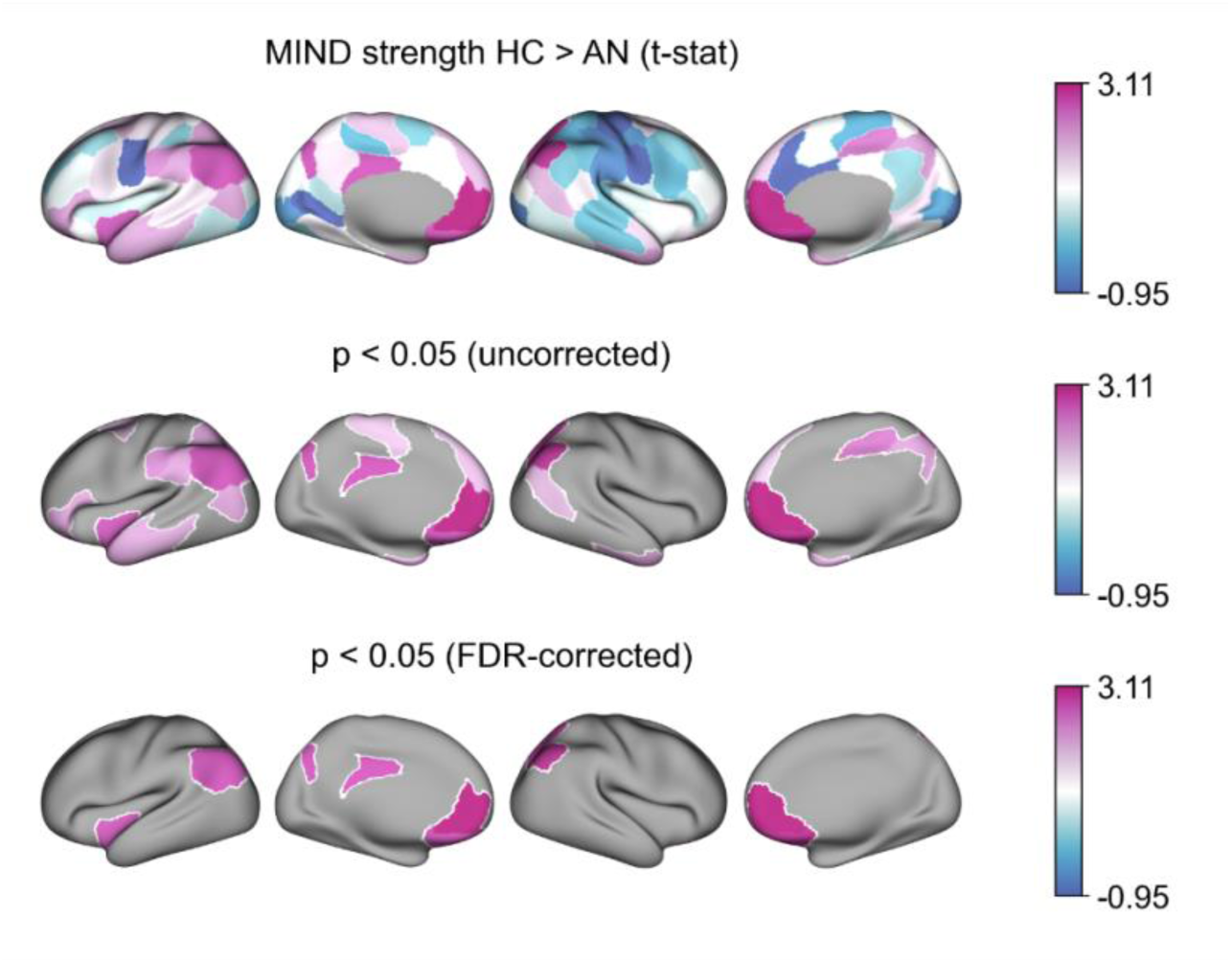
Region-wise differences in MIND connectivity between patients and controls. The upper panel shows the spatial distribution of t-statistics from GLMs testing for higher connectivity in healthy controls compared with patients with AN, controlling for TIV and age. The middle panel highlights regions of interest (ROIs) showing nominally significant group differences (uncorrected p < 0.05). The lower panel displays the ROIs that survived false discovery rate (FDR) correction, together with their corresponding empirical t-statistic values.

**Figure 3.**
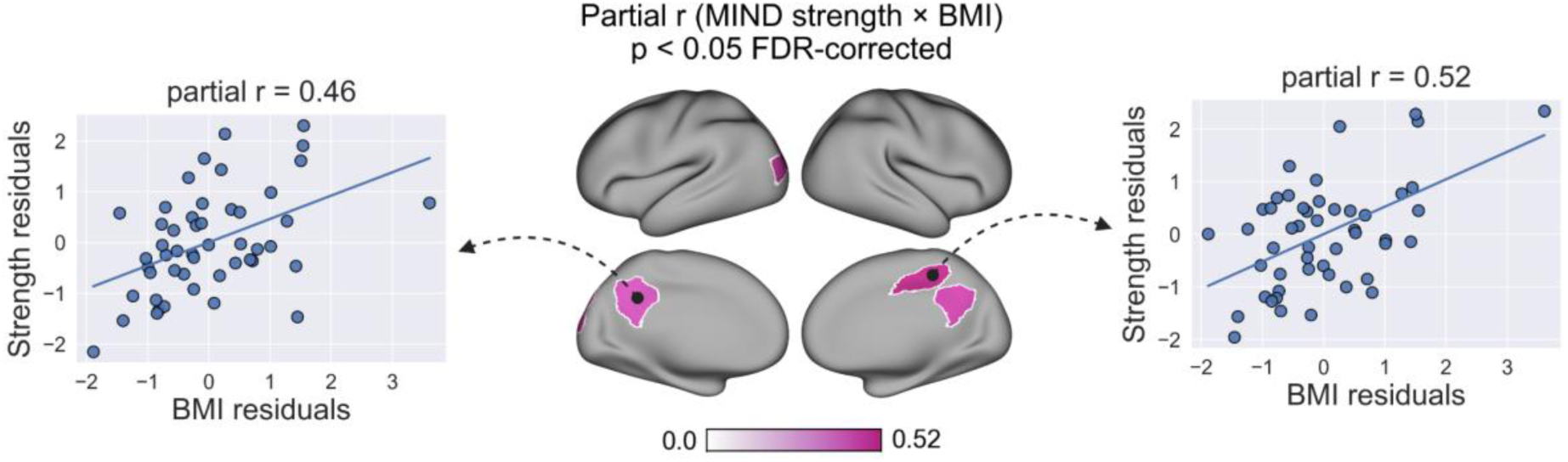
Region-wise associations between MIND connectivity strength and BMI in AN. The map shows regions where node strength was significantly associated with BMI within the AN group after FDR correction. Two exemplary scatterplots illustrate the partial correlations between BMI and MIND connectivity strength in two representative regions (values are residualized for age and TIV).

### 1.4 Regional case-control differences correlate with reward and appetite-related neurotransmitters

We next tested whether the spatial distribution of regional MIND connectivity loss in AN aligned with the density of specific neurotransmitter systems. We correlated both the uncorrected HC > AN *t*-statistic map of regional MIND strength and the map of partial correlations between regional MIND strength and BMI within the AN group with 18 cortical PET receptor/transporter atlases spanning major neuromodulatory systems (Hansen et al. 2022). Statistical significance of each correlation was assessed using 10,000 null maps that preserved the spatial autocorrelation structure of the empirical data (BrainSMASH, Burt et al. 2020) and correcting for multiple comparisons with FDR. Regions showing reduced MIND connectivity in the AN group were most strongly associated with densities of 5-HT4 (r = 0.40), 5-HT1A (r = 0.37), CB1 (r = 0.36), μ-opioid (r = 0.35), and dopaminergic D2/3 (r = 0.29; all p < 0.05; FDR-corrected). No significant associations were observed for the BMI-related map (Figure 4).

**Figure 4.**
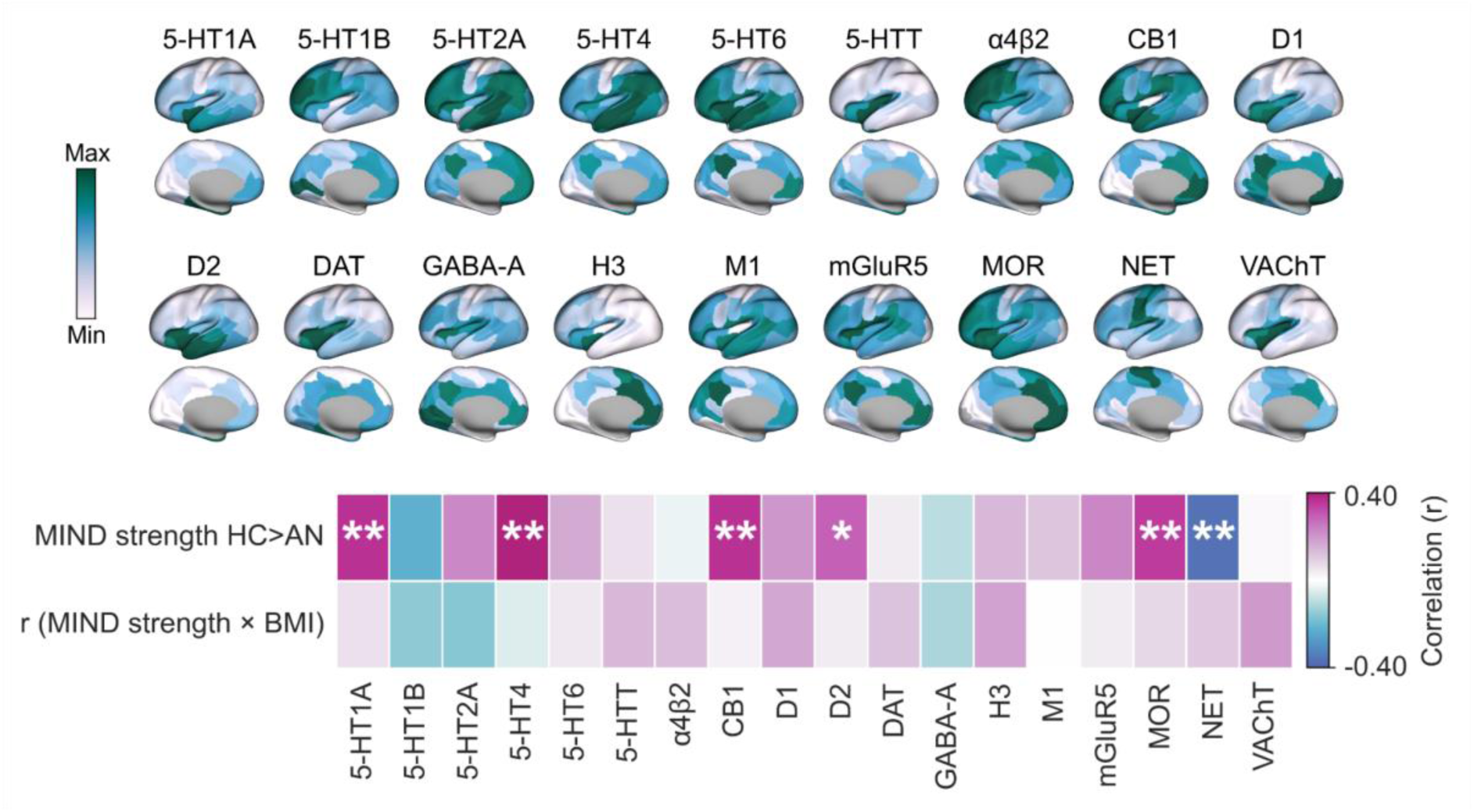
Spatial correspondence between regional MIND alterations and neurotransmitter system densities. Cortical PET receptor/transporter density maps from Hansen et al. (2022) are shown together with the spatial correlations between each neurotransmitter system and (i) the regional *t*-statistic map contrasting healthy controls with patients with anorexia nervosa (HC > AN) and (ii) the regional partial correlations between MIND strength and BMI within the AN group. Analyses were performed using the uncorrected version of both maps. Statistical significance was assessed using 10,000 null maps preserving spatial autocorrelation generated using BrainSMASH. * indicates *p* < 0.05; ** indicates *p* < 0.01 (FDR-corrected).

### 1.5 Network-driven effects on regional alterations of structural covariance

We examined whether the regional loss of MIND connectivity in AN was amplified in structurally connected regions, indicative of network-driven effects. For each brain region we computed the mean t-statistic (i.e., the HC>AN contrast) across its structurally connected neighbours and correlated this with the region’s own t-statistic. Structural neighbourhoods were defined using a template structural connectivity matrix derived from probabilistic tractography on diffusion-weighted imaging data from the healthy control group. Consistent with previous studies (e.g. Xie et al. 2025), only healthy subjects were used to build the template to provide a prototypical connectome prior to disease onset. Regions showing stronger alterations were significantly associated with greater alterations in their structurally connected neighbours (*r* = 0.42), indicating that greater MIND connectivity loss in one area was accompanied by greater loss in its structural connectivity neighbourhood (Figure 5). Two independent null models confirmed that this correlation could not be explained by spatial autocorrelation (*p*=0.0001) or by the brain’s geometric embedding, i.e., the connectivity-distance relationship (*p* = 0.005).

**Figure 5.**
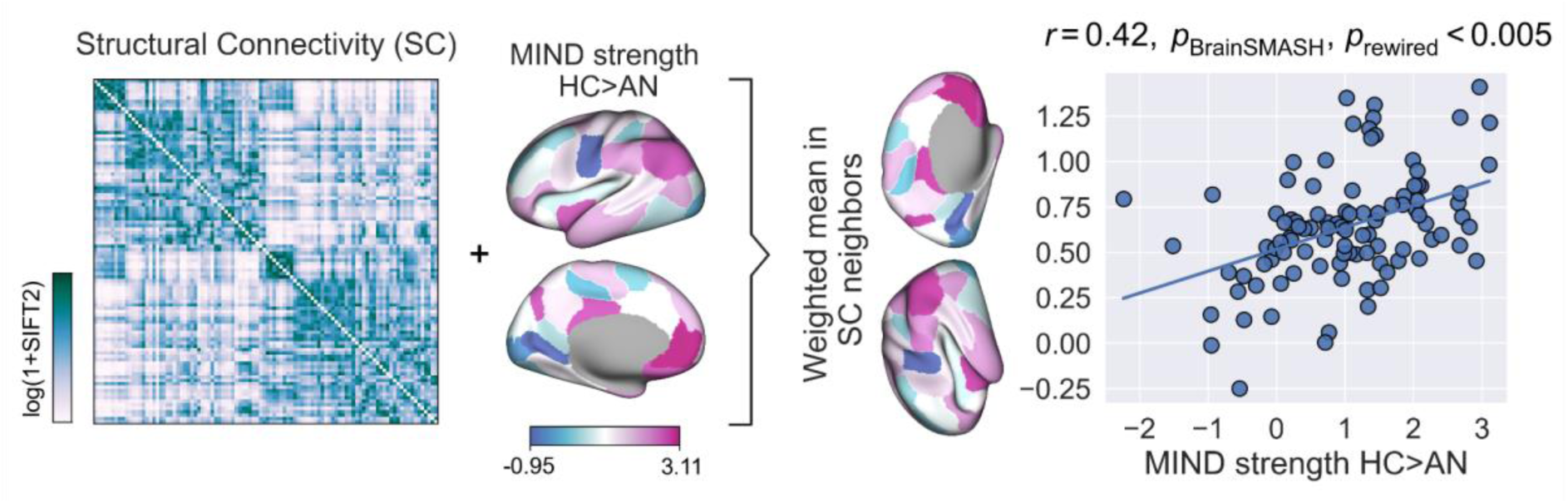
Network-driven effects on regional loss of structural covariance. Relationship between regional MIND connectivity loss in AN and the average loss observed in structurally connected neighbours, as defined by the template structural connectivity matrix derived from healthy controls. The two measures were positively correlated (r = 0.42), indicating that regions with greater MIND connectivity loss also showed greater loss in their structural connectivity neighbourhood. Statistical significance was assessed using 10,000 null maps preserving spatial autocorrelation (p_BrainSMASH_=0.0001) and a geometric null model preserving degree and edge-length distributions (p_rewired_=0.005).

### 1.6 Network-based statistical analyses of MIND connectivity in AN

Lastly, we applied a network-based statistics (NBS; Zalesky et al., 2010) approach to test for edge-wise differences between individuals with AN and healthy controls, and to relate edge-level MIND connectivity to BMI as an index of disease severity. In both analyses, age and TIV were regressed out before running NBS. Group differences were assessed with t-tests and associations with BMI using a correlational approach (see Methods). For the group comparison, we applied the NBS approach with a stringent edge-wise threshold (p < 0.001) and found one significant component comprising 12 suprathreshold connections (p = 0.025, NBS-corrected, Figure 6a). Edges in this component showed consistently higher structural covariance in healthy controls than in AN (mean t = 3.57, median = 3.53, range = 3.41–3.91). The component mainly involved connections between DAN and limbic regions, with additional links to DMN, VAN and FPN systems, suggesting a selective reduction of long-range connectivity between attentional and limbic–prefrontal circuits in AN. For the correlational analysis, we used the NBS with a stringent edge-wise threshold (p < 0.001) to isolate components of structural covariance correlated with the BMI. This analysis revealed a single significant component comprising 32 suprathreshold connections (p = 0.022, NBS-corrected, Figure 6b). Within this component, correlations with BMI were consistently positive (mean r = 0.47, median = 0.45, range = 0.43–0.55); because age and TIV had been regressed out beforehand, these values represent associations between edge-level structural covariance and BMI after accounting for these covariates. The component was dominated by long-range connections linking limbic and orbitofrontal regions with visual, VAN and DMN areas, indicating that lower BMI was associated with a loss of structural covariance mainly along inter-network pathways.

**Figure 6.**
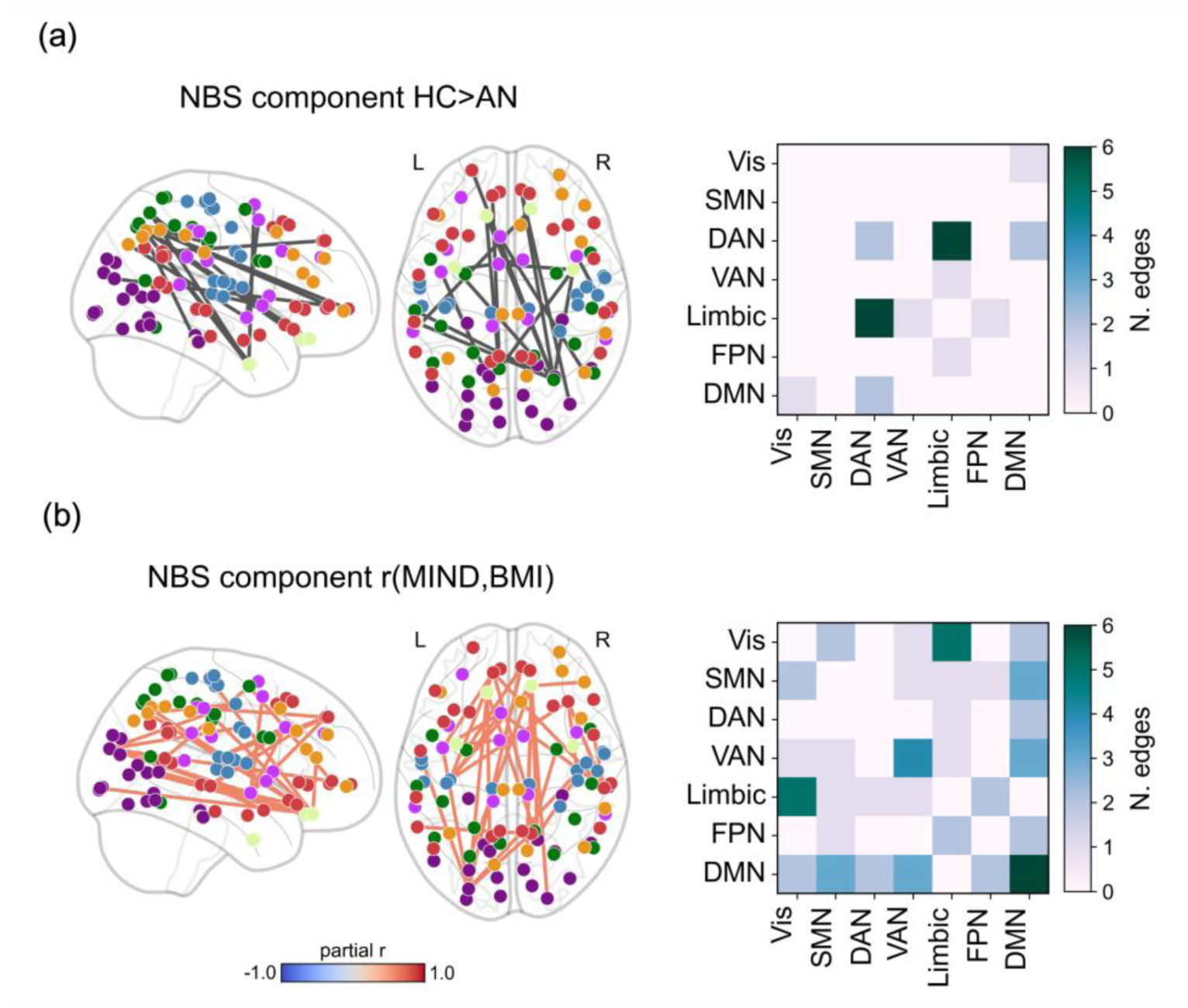
Results of the network-based statistics (NBS) analyses. (a) The left panel displays the edges of the MIND matrix showing significantly higher values in the healthy control group compared with individuals with AN, overlaid on a glass brain. The right panel depicts the distribution of these significant edges across network pairs; diagonal cells indicate counts of connections between regions within the same network. (b) As in panel (a), the left panel shows the edges of the MIND matrix, but here represents connections whose strength was significantly and positively associated with BMI (used as an index of disease severity) within the AN group. Connections are color-coded according to the magnitude of the relationship (partial r). The right panel illustrates, as above, how these significant edges are distributed across network pairs.

## Discussion

In the present study, we investigated cortical organization in AN by applying a multivariate morphometric similarity framework (MIND) that integrates multiple structural features within a network-based architecture. We first quantified group differences in MIND between patients and healthy controls to assess global and regional disruptions of cortical integrity. We then explored how these alterations relate to nutritional status and whether they align with the structural connectome and with neurotransmitter receptor topographies, thereby providing a biologically grounded model of cortical vulnerability in AN. Rather than reflecting isolated regional deviations, these findings indicate a reconfiguration of morphometric relationships across the cortical mantle that is consistent with a loss of coordinated structural integrity within the connectome. In this context, the global reduction of MIND strength, together with its association with body mass index, supports the possibility that large-scale morphometric coordination is sensitive to metabolic constraints imposed by undernutrition. While the cross-sectional design of the study precludes inferences on state dependency of these alterations, our results provide, to our knowledge, the first demonstration in AN of a global decrease in multivariate morphometric similarity that scales with nutritional status, extending prior evidence focused on univariate cortical metrics (Collantoni et al. 2024; De La Cruz et al. 2021). This interpretation is coherent with previous longitudinal morphometric work showing partial normalization of cortical thickness and surface-based measures with weight restoration, suggesting a general susceptibility of cortical architecture to energetic stress. Future longitudinal work should determine whether MIND can capture neurostructural recovery during refeeding and how these changes relate to clinical recovery.

The loss of morphometric similarity was not uniformly distributed but selectively involved transmodal cortical networks, including the limbic, default mode, frontoparietal, and dorsal attention systems. These networks lie at the apex of the cortical hierarchy, integrating sensory, cognitive, and affective information across domains (Margulies et al. 2016). Their architecture is distinguished by long-range connectivity, high synaptic density, and elevated metabolic demand, properties that confer high computational capacity but also increased vulnerability to energetic or neurochemical stress (Crossley et al. 2014). The preferential disruption of these high-order systems accords with the principle of hub vulnerability, whereby metabolically costly, high-centrality nodes are disproportionately affected under sustained energetic constraints. This topographic pattern offers a plausible network-level substrate for the cognitive rigidity, excessive self-monitoring, and emotional overcontrol that typify AN, and is consistent with prior rs-fMRI evidence implicating DMN and FPN dysconnectivity in acute AN and in relation to clinical features and treatment response, as well as with longitudinal work showing persistent network disruption despite weight restoration (Boehm et al. 2014; Kaufmann et al. 2023).

Region-wise analysis paralleled the global and network-based findings, with FDR-surviving nodes predominantly located in DMN and limbic areas. Additional BMI-strength associations emerged in visual, ventral-attention and default mode regions, indicating that morphometric similarity loss also affects areas involved in attentional and visual-perceptual processing. This pattern may align with evidence that AN is characterized by altered visual alterations and atypical processing of body-related stimuli within occipitotemporal and parietal cortices (Madsen et al. 2013; Suchan et al. 2013). Network-based statistics provided convergent evidence for these morphometric similarity findings across analytical scales. The most affected connections in AN involved long-range interactions between dorsal attention and limbic regions, extending toward default mode, ventral attention, and frontoparietal systems. Within the patients group, higher BMI was associated with stronger coupling across partially overlapping networks linking limbic and orbitofrontal areas with visual and attentional systems. These findings further support the hypothesis that undernutrition primarily disrupts large-scale integrative communication across transmodal networks, while connectivity within primary sensory systems remains relatively preserved. At the mesoscale, alterations followed the topology of the structural connectome: regions losing morphometric similarity tended to cluster along their anatomical neighbors, consistent with models of co-vulnerability and network propagation (Seeley et al. 2009). It is plausible that local perturbations in microstructural integrity, potentially related to altered energy availability, spread with interconnected network neighborhoods, leading to a progressive loss of morphometric coherence across densely linked cortical hubs.

The spatial distribution of MIND loss further aligned with receptor density gradients for serotonergic (5-HT₁A, 5-HT₄), dopaminergic (D₂/D₃), cannabinoid (CB₁), and μ-opioid (MOR) systems. These neuromodulatory pathways regulate appetite, reward valuation, emotional control, and interoception, domains that are altered in AN (De Vry & Schreiber 2000; Himmerich & Treasure 2018; Kantonen et al. 2021). Prior PET studies have reported altered serotonergic tone, dopaminergic signaling, and dysregulation of endocannabinoid and opioid systems implicated in hedonic and interoceptive processing (Bailer et al. 2010; Miranda-Olivos et al. 2023; Pak et al. 2025). The spatial correspondence between receptor gradients and morphometric disorganization suggests that cortical vulnerability in AN follows neurochemical axes supporting these functions. This receptor–MIND concordance highlights the value of multimodal frameworks integrating morphometric connectivity with molecular and receptor-level architecture to clarify the biological basis of cortical network disruption in AN. Collectively, these findings may align with a metabo-connectomic framework for understanding cortical disorganization in AN. Within this framework, the integrity of large-scale cortical networks depends on the interplay between structural connectivity, energetic sufficiency, and receptor-mediated signaling complexity. Transmodal regions, which combine high metabolic expenditure with dense neuromodulatory input, may thus represent critical nodes where energy scarcity and altered neurotransmission jointly undermine cortical architecture. Conversely, the relative preservation of primary sensorimotor and visual systems underscores the specificity of this process, suggesting that morphometric disruption reflects functional specialization rather than diffuse cortical atrophy.

## Limitations

Several limitations should be acknowledged. First, the sample size was modest, which may have reduced statistical power and limited the detection of subtle effects. Replication in larger and independent cohorts will be important to confirm the robustness and generalizability of the present findings. Second, only female participants were included, preventing inferences about potential sex differences in cortical organization and limiting the applicability of the results to the broader population. Third, patients and controls were not perfectly age-matched, with the patient group being slightly younger; however, given the limited age range of the sample, age was included as a covariate but did not account for significant variance in any of the statistical models. Lastly, the study design was cross-sectional, which precludes conclusions about causality or the temporal evolution of morphometric alterations. Longitudinal assessments will be necessary to determine whether these abnormalities normalize with weight restoration or persist as potential trait features.

## Conclusions

This study demonstrates that cortical organization in AN is disrupted at a large scale and closely related to nutritional status. By applying a multivariate morphometric framework, it captures widespread alterations in cortical connectivity, providing new insight into the structural architecture underlying this disorder.

## Methods

### Study participants and imaging data

Patients with AN were recruited from the Eating Disorder Center of the Hospital of Padova, Italy, and all met the current diagnostic criteria for AN according to the DSM-5 (DSM-5). Healthy controls (HC) were recruited from the general population through contact with the experimenters. Exclusion criteria for both groups were: (1) age under 14 years; (2) male gender; (3) presence of significant neurological comorbidities; (4) diagnosis of psychosis or substance use disorders; (5) history of hypomanic or manic episodes; and (6) known contraindications to MRI: metallic or electronic implants, pacemakers, cochlear implants. Additional exclusion criteria for HC were the presence of a current or lifetime diagnosis of an eating disorder, as assessed with the Eating Disorders section of the Structured Clinical Interview for DSM-5 Disorders (SCID-5). Among patients with AN, 26 were taking psychotropic medications (20 antidepressants, 12 antipsychotics, 4 benzodiazepines), whereas none of the healthy controls were taking any psychotropic medication. The study was approved by the Ethics Committee of the Hospital of Padova. Written informed consent was obtained from all participants prior to enrollment. In the case of underage participants, consent was provided by their parents or legal guardian.

MRI data were collected during the same experimental session following standardized acquisition protocols. All scans were performed on a 3 T Philips scanner equipped with a 32-channel head coil. High-resolution T1-weighted anatomical images were acquired using a 3D turbo field echo (TFE) sequence with SENSE parallel imaging (TR/TE = 6.7/3.1 ms; flip angle = 8°; voxel size = 1 mm isotropic; FOV = 240 × 240 × 181 mm). Diffusion-weighted data were obtained using a single-shot spin-echo echo-planar imaging (EPI) sequence with multiband and SENSE acceleration (TR/TE = 3700/104 ms; voxel size = 2 × 2 × 2 mm³; 78 slices). The protocol comprised 116 volumes, including 12 non-diffusion-weighted (b = 0 s/mm²) images and diffusion weighting at b = 300 s/mm² (7 directions), b = 1000 s/mm² (33 directions), and b = 2000 s/mm² (64 directions). An additional set of 12 b = 0 images with reversed phase-encoding polarity (AP/PA) was acquired for distortion correction. The imaging protocol also included FLAIR, pseudo-continuous arterial spin labeling (pCASL), and functional MRI sequences, which were not analyzed in the present study.

### T1-weighted preprocessing and surface reconstruction

All T1-weighted images were processed with FreeSurfer (version 7.4.1; https://surfer.nmr.mgh.harvard.edu; Fischl 2012) using the *recon-all* pipeline with default parameters. The quality of skull stripping, tissue segmentations and surface reconstructions was visually inspected, and manual edits were applied when necessary. No subject in the present cohort failed reconstruction or required manual corrections.

### Construction of MIND networks

To build structural similarity networks, cortical thickness (CT), mean curvature (MC), volume (Vol), sulcal depth (SD) and surface area (SA) were extracted at each cortical vertex. After applying the cortical parcellation, each ROI was represented by the distributions of these features across its constituent vertices. MIND (Sebenius et al. 2023) quantifies similarity between regions by computing a multivariate Kullback–Leibler (KL) divergence between their feature distributions, estimated via a k-nearest-neighbour approximation to improve efficiency and reduce free parameters. This divergence is inverted and rescaled so that larger divergences correspond to lower similarity scores, yielding values between 0 and 1. In the resulting MIND matrix, higher values reflect greater morphometric similarity between regions. Full mathematical details are provided in Sebenius et al. (2023).

### DWI preprocessing

We visually inspected DWI data for motion-related artefacts and slice-wise instabilities following consensus recommendations, and we excluded volumes with gross artefacts from further analysis. We denoised the data by using the Marchenko - Pastur PCA (Veraart et al. 2016), and we removed Gibbs ringing artefacts with a subvoxel-shift–based approach (Kellner et al. 2016). We then corrected the data for susceptibility-induced distortion, motion and eddy currents with a single-step resampling procedure implemented in in the *dwifslpreproc* function of MRtrix3 (Andersson & Sotiropoulos 2016; Tournier et al. 2019). We applied biasfield correction using the N4 algorithm from Advanced Normalization Tools (ANTs; Tustison et al. 2010). We skull-stripped the mean *b0* image with SynthStrip (Hoopes et al. 2022) and rigidly registered it to the high-resolution T1-weighted image using FSL (cost function: normalized mutual information). We visually assessed the accuracy of each registration. Finally, we applied the transformation to the DWI header (without regridding) while ensuring appropriate rotation of the diffusion gradient table.

### Tractography and connectome generation

We performed whole-brain probabilistic tractography and structural connectivity mapping on DWI data aligned to anatomical space. We obtained tissue segmentations for anatomically constrained tractography (ACT; Smith et al. 2012) using Hybrid Surface–Volume Segmentation (HSVS; Smith et al. 2020). We estimated fiber orientation distributions (FODs) using multi-shell, multi-tissue constrained spherical deconvolution (Jeurissen et al. 2014). We averaged white matter (WM), grey matter (GM) and cerebrospinal fluid (CSF) response functions across participants to generate group-consensus, unbiased response functions. We then generated ten million fibers, whole-brain tractograms with the iFOD2 probabilistic tracking algorithm (Tournier et al. 2010) using dynamic WM seeding. We reduced streamline density biases and enhanced biological plausibility with the SIFT2 algorithm (Smith et al. 2015). We then constructed weighted and symmetric structural connectivity matrices based on the Schaefer 100 regions 7 Networks atlas (Schaefer et al. 2018). Finally, we generated a group consensus structural connectivity matrix using a distance-dependent thresholding approach (Betzel et al. 2019).

### Statistical analysis of structural covariance networks

#### Generalized linear models (GLMs)

We used GLMs to assess global, network-specific and node-specific differences in MIND strength between subjects with AN and healthy controls. Each GLM included age and estimated total intracranial volume (TIV) as covariates. We did not control for the effect of sex, since all study participants were female. When we assessed across-subject associations (e.g. BMI and MIND strength relationships), we complemented the GLM analysis with the partial correlations between variables net of age and VIT. In each analysis, we corrected for multiple comparisons using the FDR correction.

#### Network-driven effects on regional alterations of structural covariance

We tested whether regional loss of MIND connectivity in AN followed a network-driven pattern. For each brain region, we first computed the average alteration across all structurally connected regions, as defined by a template structural connectivity matrix built from probabilistic tractography on the DWI data of the healthy control group. This procedure yielded, for every region, a pair consisting of its alteration value and the mean alteration of its structural neighbours. We then quantified the correspondence between these two measures by computing a correlation coefficient. We evaluated the statistical significance of the correlation coefficient using two null models. As the first null model, we used the variogram-matching approach of Burt et al. (2020), as implemented in the Python package BrainSMASH, to generate 10,000 surrogate maps with the same spatial autocorrelation as the empirical map. Full details of the method and its validation are provided in Burt et al. (2020). The second null model controls for the effect of network-geometry, i.e., the relationship between connectivity and distance. In practice, we generated N = 1000 degree- and edge length-preserving random structural connectivity matrices. For the full details we refer to the paper that first described the null model (Betzel & Basset 2018). Null model instances were generated using the python tool *netneurotools (*https://netneurotools.readthedocs.io/en/latest/index.html).

#### Network-based statistics (NBS)

We applied the Network-Based Statistic (NBS; Zalesky et al., 2010) to identify subnetworks showing altered structural covariance between individuals with AN and healthy controls and to detect subnetworks whose edge-wise connectivity correlated with BMI within the AN group. Before running NBS, age and estimated total intracranial volume were regressed out from each edge of the structural covariance matrices. For the correlational analysis, age and estimated total intracranial volume were also regressed out from BMI so that associations were estimated on residual values free of these effects. For the group contrast, edge-wise two-sample t-tests were performed on the residual connectivity matrices of the two groups. For the BMI analysis, the correlational version of NBS was used to assess associations between edge-wise connectivity and BMI residuals within the AN group. We used a stringent primary threshold equivalent to p < 0.001 to define suprathreshold connections. The size of each connected component was compared with an empirical null distribution generated by 5,000 random permutations to obtain component-level corrected p-values, with significance set at p < 0.05 at the component level. All NBS analyses were implemented in Python using the brainconn package (https://brainconn.readthedocs.io/en/latest/index.html) together with publicly available code for the correlational NBS framework (https://github.com/GidLev/NBS-correlation; Levakov et al. 2021).

### PET/SPECT receptor atlases

We utilized a curated dataset of PET/SPECT receptor and transporter atlases to contextualize our results (Hansen et al., 2022). This dataset comprises regional measures and/or semiquantitative proxies of receptor and transporter densities for 18 different targets, spanning the following neurotransmitter systems: glutamate (mGluR5), GABA (GABAA), serotonin (5HT1A, 5HT2A, 5HT1B, 5HT6, 5HT4, 5HTT), acetylcholine (A4B2, VAChT, M1), noradrenaline (NAT), histamine (H3), opioid (MU), cannabinoid (CB1), and dopamine (D1, D2, DAT). We downloaded the atlases from a public repository (https://github.com/netneurolab/hansen_receptors) associated with the publication by Hansen et al. (2022). To assess the statistical significance while accounting for spatial autocorrelation we use the BrainSMASH approach (N.Nulls =10,000).

## Data and code availability statement

Data supporting the findings of this study are not publicly available due to privacy and ethical restrictions, but are available from the corresponding author upon reasonable request. Custom analysis code is also available upon reasonable request.

## Author Contributions

MF: Conceptualization, Methodology, Formal Analysis, Writing - original draft

VM: Data curation, Resources, Writing - original draft

SG: Data curation

AB: Methodology, Writing - review & editing

RM: Data curation, Resources, Writing - review & editing

AF: Methodology, Resources, Writing - review & editing

EC: Conceptualization, Methodology, Supervision, Project administration, Writing - original draft

## Data Availability

All data produced in the present study are available upon reasonable request to the authors

