## Supplementary Materials for "Local-Global breakdown of Cortical Similarity Networks in Anorexia Nervosa"

| **Region** | **Network** | **t(HC>AN)** | **pval - FDR** |
| --- | --- | --- | --- |
| SalVentAttn_2 | SalVentAttn | 2,650 | 0,048 |
| Limbic_1 | Limbic | 2,685 | 0,048 |
| Cont_3 | Cont | 2,674 | 0,048 |
| Cont_4 | Cont | 2,711 | 0,048 |
| Default_4 | Default | 2,685 | 0,048 |
| Default_7 | Default | 3,111 | 0,048 |
| DorsAttn_12 | DorsAttn | 2,818 | 0,048 |
| Limbic_4 | Limbic | 2,963 | 0,048 |
| Cont_6 | Cont | 2,917 | 0,048 |
| Default_20 | Default | 3,109 | 0,048 |

**Table T1.** Significant differences in regional MIND strength between HC and patients with AN net of age and TIV effects.

| **Region** | **Network** | **partial r** | **pval - FDR** |
| --- | --- | --- | --- |
| Vis_8 | Vis | 0,482 | 0,023 |
| Default_13 | Default | 0,431 | 0,049 |
| SalVentAttn_11 | SalVentAttn | 0,521 | 0,012 |
| Default_24 | Default | 0,460 | 0,030 |

**Table T2.** Significant partial correlations between regional MIND strength and BMI in the AN group net of age and TIV effects.
